# Incidence of Deep Venous Thrombosis Post Cardiac Surgery: A Prospective Observational Duplex Venous Screening

**DOI:** 10.1101/2025.03.05.25323437

**Authors:** Abdul Hasan Siddiqui, Masood A Shariff, Kimberly C Bowman, Jennifer Musumeci, Kristen Roy, Ahmed Klobocista, William Molloy, Theodore J Maniatis, Joseph T McGinn, Kuldeep Singh, John P Nabagiez

## Abstract

**Background:** Prior studies investigating deep vein thrombosis (DVT) post cardiac surgery have included mostly on pump cardiac surgery. We aimed to evaluate the frequency of DVT in consecutive series of patients undergoing cardiac surgery by sequential Duplex Venous Sonography (DVS).

**Methods:** A single center prospective cohort study conducted on patients undergoing cardiac surgery from 2012 to 2014. All patients underwent sequential DVS at three-time intervals: Preoperatively, Day of Discharge and 5 weeks Postoperatively. A subgroup analysis was done to compare DVT and non-DVT groups to look for possible predictors for development of DVT.

**Results:** 124 patients underwent screening DVS. 77.4% underwent off-pump CABG, 2.4% on-pump CABG, and 20.1% valve surgery. Positive asymptomatic DVS was observed in 11 of 124 eligible patients (8.9%). Off-pump CABG (10 of 96; 10%) and valve replacement/repair (1 of 25; 4%). Mean age of the DVT and non-DVT groups was 65.5±9.3 and 66.0±10.2 and females were 18% and 29%, respectively. The groups did not differ in surgical time (3.9±1.0 vs. 3.8±1.0 hours), intubation time (14.9±7.0 and 12.1±5.3 hours) and length of stay (6.7±5.0 and 6.6±5.4 days), DVT and non-DVT groups, respectively. Patients with DVT had a higher requirement for blood transfusions (55% vs. 20% [P<0.05]). Thirty-day hospital readmissions were 9% for DVT versus 13% for non-DVT. All patients received thromboprophylaxis and physical therapy postoperatively.

**Conclusions:** To our knowledge, first of its kind, a prospective, observational DVS study to evaluate DVT in Cardiac Surgery patients. Despite optimal thromboprophylaxis, 9% of cardiac surgery patients develop otherwise asymptomatic DVT when off-pump CABG is performed. Off pump CABG may have better outcomes with respect to postoperative DVT. We found thromboprophylaxis combined with physical therapy to be safe and effective to prevent symptomatic DVT, whereas routine preoperative screening did not influence the results in our study. Despite limited sample size, we found the DVT group included a higher percentage of patients who smoked, required blood transfusion and experienced prolonged intubation or reintubation.

## INTRODUCTION

Venous Thromboembolism is a common major illness seen in hospitalized patients resulting in increased morbidity and mortality (1). Postoperative deep vein thrombosis (DVT) is one of the quality indicators related to surgical outcomes (2). Most venous thrombosis episodes go clinically unnoticed. When venography or radiolabeled fibrinogen is used in asymptomatic postoperative patients, more than 20% of them are found to have evidence of DVT (3). Studies related to DVT in cardiac surgery patients are limited and no clear-cut guidelines exist for thromboprophylaxis and screening for DVT in the current literature (3,4). The prevalence of DVT post cardiac surgery detected by duplex ultrasound varies between 0.7% and 48% (5–7). In mostly retrospective studies, the incidence of symptomatic venous thromboembolism ranged from 0.5% to 3.9%; and fatal and nonfatal pulmonary embolisms, up to nearly 4% (8). The guidelines for the prevention and treatment of DVT, although well established with respect to other surgeries, are very limited related to cardiac surgery due to lack of strong evidence. In a collective review on prevention of venous thromboembolism (8), no specific recommendations were made for cardiac surgery patients undergoing coronary artery bypass grafting (CABG), suggesting that DVT prevention has never been seen as a priority by cardiac surgeons due to the administration of large amount of heparin intraoperatively as well as the antithrombotic effects of procedures utilizing cardiopulmonary bypass (CPB) (5,9–12). In fact, the recommendations are inconsistent with European Association for Cardio-Thoracic Surgery (EACTS) recommending DVT prophylaxis with heparin for all post cardiac surgery whereas American College of Clinical Pharmacy (ACCP) guidelines recommends against routine anticoagulation post cardiac surgery due to an elevated risk of hemorrhagic pericardial effusion and tamponade (13,14). However, these are merely grade 2 recommendations and based on poor evidence (15). Hence it is important to conduct a study to evaluate the true incidence of DVT and to establish predictors and risk factors for the development of DVT in patients undergoing cardiac surgery.

Because off-pump CABG (OPCAB) avoids the use of CPB and cardiac arrest, the level of anticoagulation used by many surgeons is generally lower than that found in the standard procedure with CPB. The CPB-related inflammatory response caused by the activation of plasma protein systems such as contact system, intrinsic and extrinsic coagulations, complement, and fibrinolytic system, is accordingly reduced (16–19). Postoperative hypercoagulability is a well-documented state after general and orthopedic operations and is responsible for significant postoperative morbidity (2,20,21). At our institution, mostly off-pump CABG surgeries are performed albeit with full heparinization as used with CPB and our goal is to evaluate the incidence of DVT in patients undergoing cardiac surgery through sequential perioperative screening by bilateral lower extremity duplex venous sonography (DVS). To our knowledge, first of its kind, a prospective, observational DVS study to evaluate DVT in Cardiac Surgery patients.

## METHODS

From January 2012 to July 2014, a single center prospective cohort study was conducted on patients undergoing cardiac surgery. The study was approved by Northwell Health’s Institutional Review Board (#SIUH11-044) and all patients gave informed consent for participation.

### Study Population

Inclusion criteria for the study included patients undergoing isolated cardiac surgery, off-pump CABG or valve (aortic or mitral valve). Exclusion criteria consisted of prior history of DVT and concomitant valve and CABG operation. The patients were prospectively recruited to participate based on availability to follow up until 5-weeks postoperatively. All the patients underwent DVS scans of the lower extremities preoperatively, at Baseline, and two additional scans postoperatively at the time of Discharge, and at 5-Week.

The following data were collected: age, sex, weight, height, preoperative hemoglobin, presence of diabetes, chronic obstructive pulmonary disease, peripheral vascular disease and risk factors for thromboembolism. The perioperative information included surgical time, intubation time, blood products, estimated blood loss, vein harvest site, cardiopulmonary bypass access site and postoperative complications. Predictors of postoperative DVT were collected: physical therapy initiation day, body mass index, cancer profile, peripheral vascular disease, chronic obstructive pulmonary disease (COPD), hypertension; current smoking; left ventricular ejection fraction, and serum creatinine. The length of stay, need for ICU re-admission and discharge disposition were reviewed.

### Surgical Procedure

The CABG patients underwent endoscopic vein harvest cauterization of branches or open dissection, the branches ligated with suture or clips. Valve surgery is performed via right thoracotomy: medial 2nd intercostal space for AVR and lateral 5th intercostal space for MVR with CPB access via femoral artery and vein or subclavian artery and femoral vein. Standard sternotomy access for CPB is accomplished via the aorta and right atrium.

Minimally invasive coronary artery bypass (MICS) is performed through a left thoracotomy in the 5th intercostal space in selected patients (19). This technique occasionally requires CPB support via femoral artery and vein on a beating heart without cardioplegic arrest.

### Venous Doppler Ultrasonography

All the patients recruited for the study were non-invasively screened for DVT using serial venous ultrasonography. The ultrasound system General Electric Company, LOGIQ E9 was equipped with 4-MHz and 7.5-MHz transducers with duplex and color Doppler capability. Protocol imaged both legs from the groin distally including the calf veins, the common femoral, deep femoral, superficial femoral and popliteal veins. Imaging of the calf veins (anterior tibial, posterior tibial, peroneal sural and soleus muscle and draining veins of the lateral and medical heads of the gastrocnemius muscle) were obtained both transversely and longitudinally and confirmed with the color flow accentuation after the lower third of the calf was squeezed.

Proximal DVT was defined as incomplete compression of common femoral, superficial femoral deep femoral or popliteal veins (with or without thrombosis of the calf vein). Isolated calf DVT was defined as loss of compressibility and loss of the expected increase in color Doppler signal with flow augmentation. All patients were screened at least three times during the study.

The noninvasive vascular studies were performed at the institution’s vascular laboratory by vascular ultrasonographers. In the event of DVT detection by noninvasive means, the patient was to be treated with full heparinization and removed from the study. Minor DVTs were defined as ‘below the knee” DVTs and the treatment decision was left at the primary physician discretion. All DVT scans were reviewed and reports collected. The relevant findings included the number and location of DVTs.

### Inpatient Postoperative Care

After surgery, patients are transferred to the cardiothoracic unit (CTU). Prolonged intubations were defined as greater than 48 hours. Physical therapy evaluation was started on all eligible patients from postoperative day one and out of bed to chair was initialed as soon as possible. Gait is evaluated thereafter based on patient’s responsiveness and hemodynamic status.

In accordance with CTU protocol, DVT prophylaxis was initiated postoperatively and continued until discharge in patients who do not require full anticoagulation for other reasons. Low-molecular-weight heparin (LMWH) or unfractionated heparin was utilized for prophylaxis.

All patients on aspirin therapy from day 1 of the surgery, using subcutaneous heparin 5000 units three times a day or enoxaparin 40 mg once a day. Intermittent pneumatic compression device was used on lower extremities bilaterally until the patient began ambulation. Prophylaxis continued if the patient was discharged to an extended care facility until discharge.

The primary end point was the detection of DVT in postoperative cardiac surgery patients. Secondary end points were factors that influenced the development of DVT: type of surgery, location, and timing of DVT, presence of pulmonary embolism, bleeding requiring thoracotomy, complications related to surgery, operation time, blood transfusion requirement and length of stay.

### Statistical analysis

Sample size was calculated based on the published incidence of thrombosis in cardiothoracic surgery population with a 95% CI for the rate of DVT of 0.7–48%. A sample size of 385 was required but the study was aborted before reaching the estimated sample size. The study was terminated due to the slow/insufficient rate of accrual and the patient admission and surgical priority dynamics changed during the Recruitment Period, as the number of electives cases dropped and In-house patients fell in the Emergency/Urgent category, limited our recruitment due to screening failures.

Preoperative and postoperative characteristics for continuous variables are reported as mean ± standard deviation (SD) and categorical variables as frequency and percentages. Binary logistic regression was used to determine the independent predictors of DVT using all the variables with *p* value less than 0.2 (univariate), with highly correlated variables excluded to avoid model confounding. Variables significant at the *p* value less than 0.05 level were retained as predictors. Statistical analysis was conducted with SPSS version 17.0 software.

## RESULTS

A total of 846 patients were screened to participate in the study, 135 consented to participate. Six subjects did not undergo surgery, and five subjects did not undergo baseline DVS. A total of 124 patients completed the study protocol (Figure 1). The surgery procedures were CABG 99 (79.8%) and valves, 25 (20.2%). CABG was 97% off-pump and 3% on-pump. The valve surgeries were aortic valve replacement (AVR) 13 (52%) and mitral valve was operated 12 (48%) (Replacement 6 and valvuloplasty 6). Patient characteristics included 89 males (71.8%) and 35 females (28.2%), mean age of 65.9±10.1 years (range 56–76). Baseline characteristics and operative and postoperative findings are summarized in Table 1.

**Figure 1:**
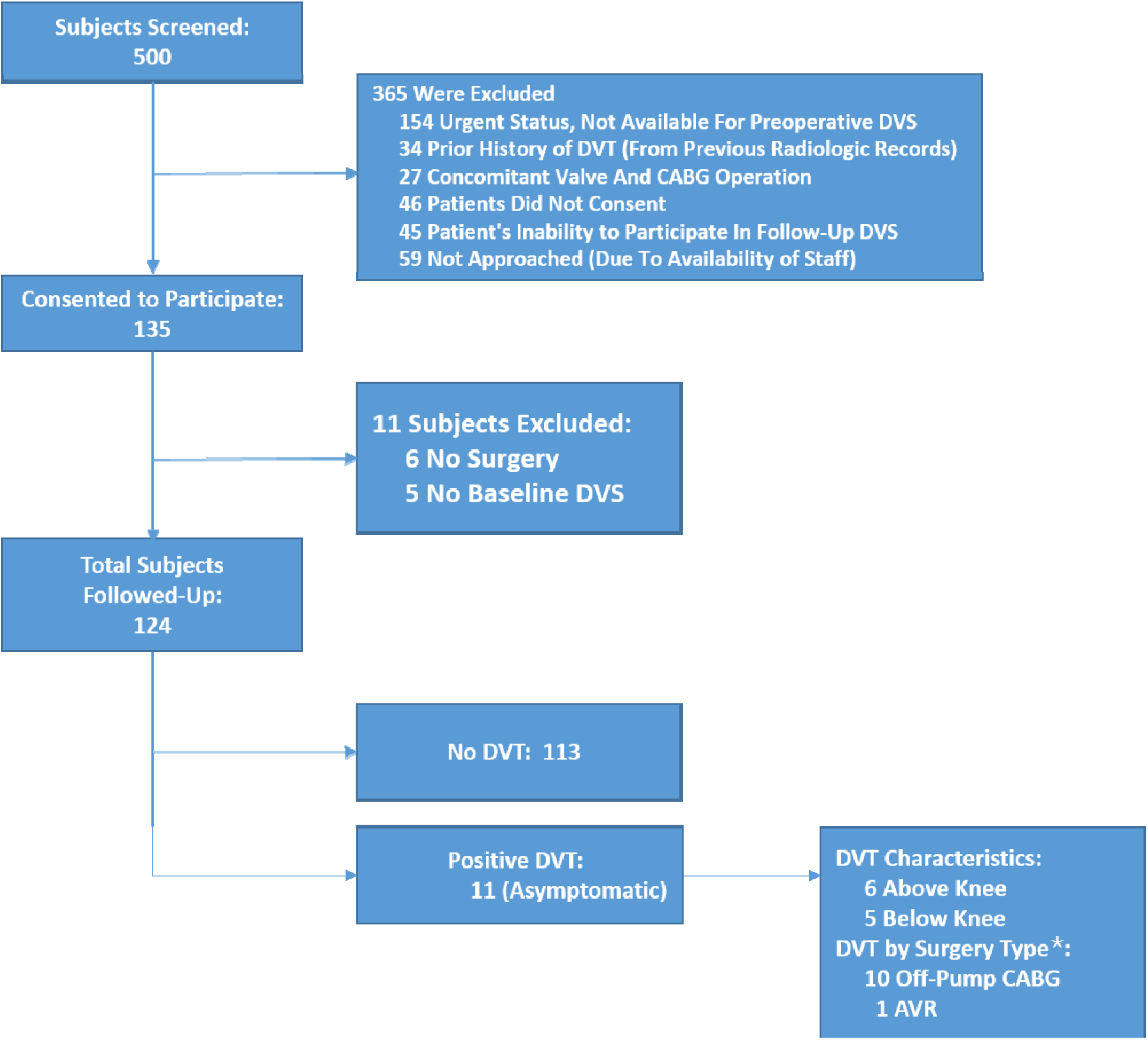
Flow diagram of the recruitment process for the study. (AVR, aortic valve replacement; CABG, coronary artery bypass grafting; DVT, deep vein thrombosis; DVS, duplex venous sonography)

**Table 1:** Patient Preoperative Risk, Operative Variables and Postoperative Outcomes.

|  |  |
| --- | --- |
| N | 124 |
| Age, year | 65.9 ± 10.1 |
| BMI (kg/m2) | 29.2 ± 5.2 |
| Risk factors: |  |
| Male | 89 (71.8%) |
| History of Smoking | 73 (58.9%) |
| Current Smoker | 28/73 (22.6%) |
| Diabetes | 52 (41.9%) |
| Hypercholesterolemia | 96 (77.4%) |
| Dialysis | 3 (2.4%) |
| Hypertension | 109 (87.9%) |
| Cerebral Vascular Disease | 14 (11.3%) |
| Chronic Obstructive Lung Disease | 23 (18.5%) |
| Peripheral Vascular Disease | 8 (6.5%) |
| Surgery Type |  |
| CABG | 99 (79.8%) |
| Off-Pump | 96/99 (97%) |
| On-Pump | 3/99 (3%) |
| Valve | 25 (20.2%) |
| Aortic | 13/25 (52%) |
| Mitral | 12/25 (48%) |
| Surgery time, hours | 3.7 ± 0.9 |
| Blood transfusion required | 29 (23%) |
| Intubation time, hours | 12.3 ± 5.5 |
| Postoperative Day of Mobilization, Median (range) | 2 (1-10) |
| Complications: |  |
| Prolonged Ventilation | 6 (5%) |
| Re-intubated During Hospital Stay | 5 (4%) |
| Renal Failure | 4 (3%) |
| Reoperation for Bleeding/Tamponade | 3 (2%) |
| Stroke | 1 (1%) |
| Transient Ischemic Attack | 1 (1%) |
| Paralysis-Transient | 1 (1%) |
| Pulmonary Embolism | 1 (1%) |
| Length of Stay, days | 6.6 ± 5.3 |
| Discharge Location |  |
| Home | 98 (79%) |
| Nursing Home | 19 (15%) |
| Rehab Center | 6 (5%) |
| Transferred to different hospital | 1 (1%) |
| Readmission, 30-days PostOP | 15 (12%) |
| Values are mean ± standard deviation or n (%). | |

Primary endpoint was 8.8% (n=11) of subjects that developed DVTs all asymptomatic (Table 2). Of these 11 DVTs, 6 (55%) were above the knee and 5 (45%) below the knee. DVT characteristics in terms of the extremity that was intervened upon for vein harvesting in CABG surgery or femoral artery/vein CPB cannulation: 8 were on ipsilateral and 3 on contralateral extremity. Nine patients had an isolated DVT, one patient had 2 DVTs and one patient had more than 3 DVTs. The patient with 3 below knee DVTs of note had bilateral saphenous vein harvesting. In terms of surgery type, CABG patients that developed DVTs were all in the off-pump group (10 out of 99, 10%) and in valve patients one DVT developed in one AVR patient (0.8%).

**Table 2:** DVT Characteristics.

|  |  |
| --- | --- |
| Deep Venous Thrombosis (DVT) | 11 (9%) |
| No. DVTs per patient |  |
| 1 | 9 |
| 2 | 1 |
| 3+ | 1 |
| DVT location: |  |
| No. Above knee | 6/11 (55%) |
| No. Below knee | 5/11 (45%) |
| Association with Vein Harvest or Femoral CPB Cannulation |  |
| Ipsilateral | 8/11 (73%) |
| Contralateral | 3/11 (27%) |
| DVT by Surgery Type: |  |
| CABG | 10 (8%) |
| Off-Pump | 10/99 (10%) |
| On-Pump | 0 |
| Valve | 1 (0.8%) |
| Aortic-Replacement | 1/25 (4%) |
| Other Findings: |  |
| Symptomatic Arm DVT | 3 (2%) |
| Seroma | 3 (2%) |
| Hematoma | 2 (2%) |
| Values are mean ± standard deviation or n (%). |  |

A subgroup comparison between the DVT and non-DVT patients is shown in Table 3. In terms of mean and standard deviation the groups were similar in age, body mass index, intubation time, surgery time and length of stay. There was a higher rate of smokers in the DVT group (73%) than in the non-DVT group (58%) (p=0.328), More patients in the DVT group underwent urgent surgery (64%) compared to elective surgery (36%) (p=0.289). There was a higher rate of blood transfusions in the DVT group, 6 out of 11 (55%), compared to the non-DVT group with a rate of 20% (p=0.008). There were no PAD patients in the DVT group. Readmission was also higher in the non-DVT group (15 out of 113, 13%) with only one in the DVT group (9%) (p=0.693) none of the patients were readmitted for symptomatic DVT. One patient was diagnosed with a pulmonary embolism with a negative DVS for DVT; and 3 (2.4%) patients had major bleeding requiring reoperation, one of them had a DVT.

**Table 3:**
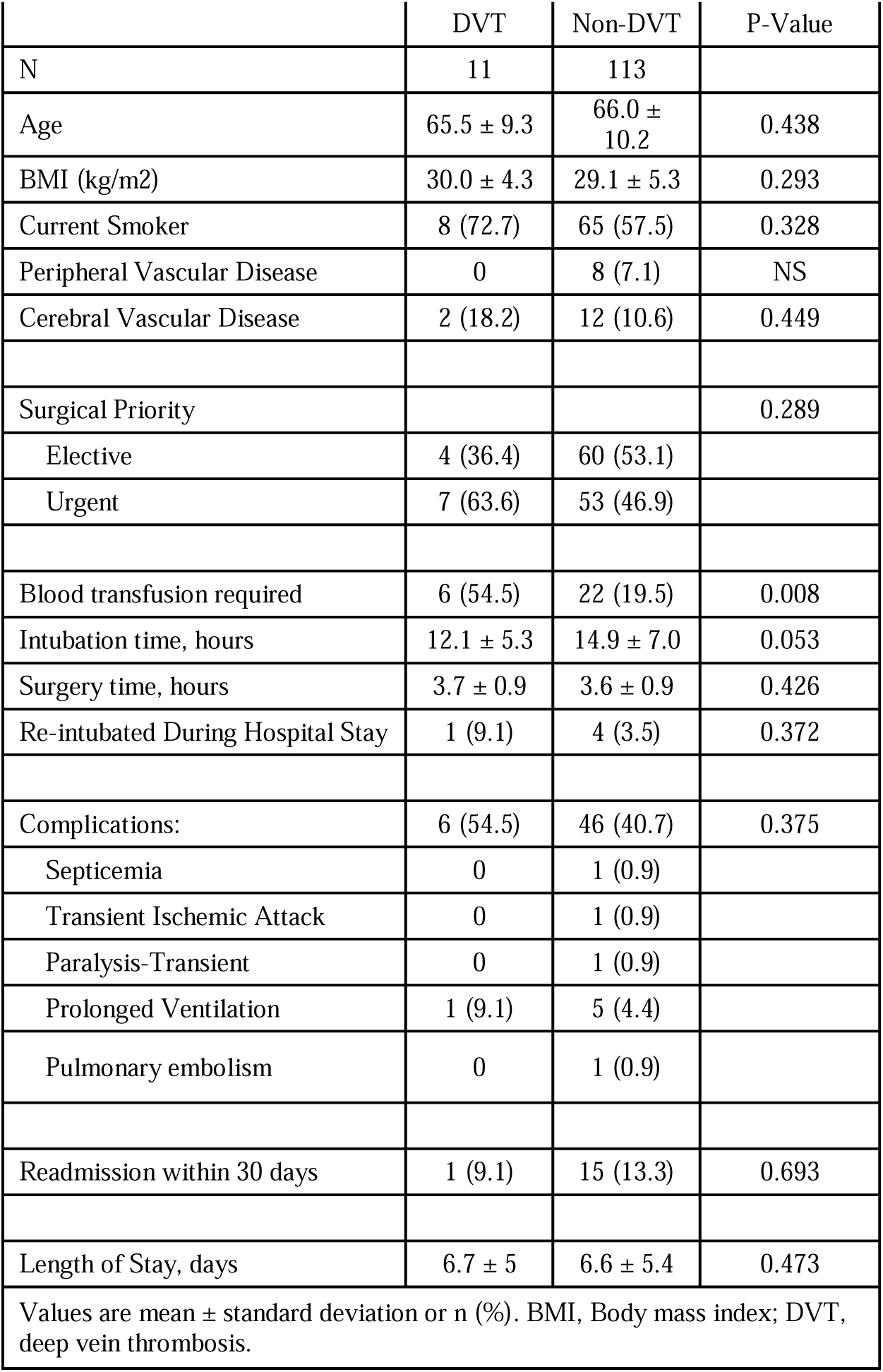
Subgroup Comparison.

|  | DVT | Non-DVT | P-Value |
| --- | --- | --- | --- |
| N | 11 | 113 |  |
| Age | 65.5 ± 9.3 | 66.0 ± 10.2 | 0.438 |
| BMI (kg/m2) | 30.0 ± 4.3 | 29.1 ± 5.3 | 0.293 |
| Current Smoker | 8 (72.7) | 65 (57.5) | 0.328 |
| Peripheral Vascular Disease | 0 | 8 (7.1) | NS |
| Cerebral Vascular Disease | 2 (18.2) | 12 (10.6) | 0.449 |
| Surgical Priority |  |  | 0.289 |
| Elective | 4 (36.4) | 60 (53.1) |  |
| Urgent | 7 (63.6) | 53 (46.9) |  |
| Blood transfusion required | 6 (54.5) | 22 (19.5) | 0.008 |
| Intubation time, hours | 12.1 ± 5.3 | 14.9 ± 7.0 | 0.053 |
| Surgery time, hours | 3.7 ± 0.9 | 3.6 ± 0.9 | 0.426 |
| Re-intubated During Hospital Stay | 1 (9.1) | 4 (3.5) | 0.372 |
| Complications: | 6 (54.5) | 46 (40.7) | 0.375 |
| Septicemia | 0 | 1 (0.9) |  |
| Transient Ischemic Attack | 0 | 1 (0.9) |  |
| Paralysis-Transient | 0 | 1 (0.9) |  |
| Prolonged Ventilation | 1 (9.1) | 5 (4.4) |  |
| Pulmonary embolism | 0 | 1 (0.9) |  |
| Readmission within 30 days | 1 (9.1) | 15 (13.3) | 0.693 |
| Length of Stay, days | 6.7 ± 5 | 6.6 ± 5.4 | 0.473 |
| Values are mean ± standard deviation or n (%). BMI, Body mass index; DVT, deep vein thrombosis. |  |  |  |

Diagnosis of DVT ranged from postoperative days 3 to 25, where 6 patients developed DVT within the first 5 postoperative days (Figure 2). Six of the patients diagnosed with above knee DVT were immediately started on anticoagulation therapy but the treatment decision of the calf DVTs were left to the primary care team for the evaluation of risk and benefits of anticoagulation.

**Figure 2:**
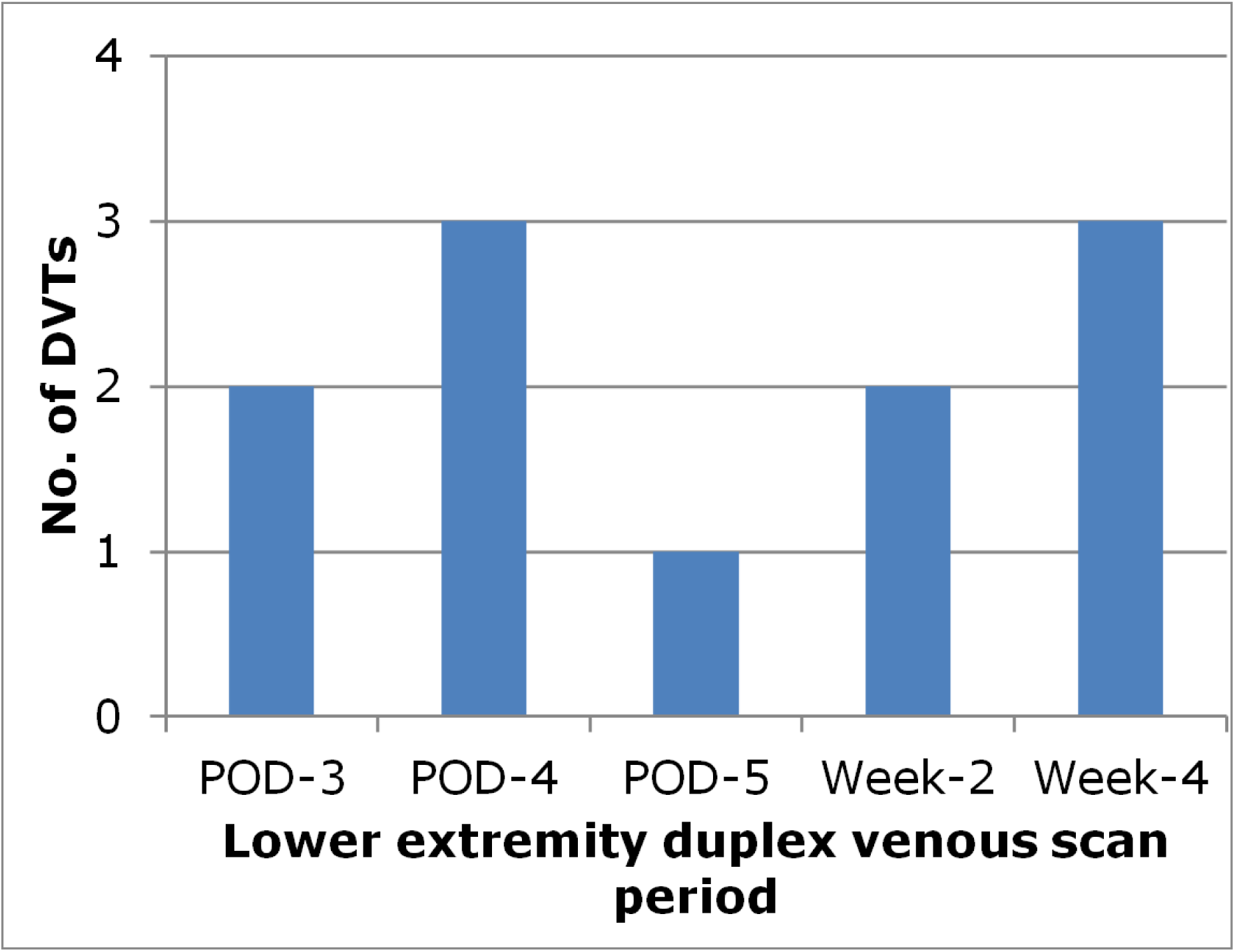
DVT Events (POD, postoperative day)

## DISCUSSION

The incidence of DVT and appropriate thromboprophylaxis related to off pump CABG is not well established in the current literature. Though inconsistent, best practice guidelines support the use of thromboprophylaxis after surgery (20–22). In our study, we prospectively screened 124 post cardiac surgery patients and the incidence of DVT was found to be 9% (11/124). As most of the patients 99/124 underwent off pump CABG surgery we conclude that the incidence of DVT in off pump CABG surgery may be as high as 10% (10/96).

There is a wide variation in the incidence of DVT reported in the published literature with respect to cardiac surgery patients (0.7 to 48%) (6). This could be explained by the variation in the methods of detection symptomatic versus surveillance, prospective versus retrospective, type of procedure, the screening method and method of thromboprophylaxis (2,22). Few prospective studies exist in cardiac surgery for detection of DVT, especially those undergoing OPCAB. Our study is unique as this is the first study, which is done prospectively on patients mostly undergoing off pump CABG surgery. Many of the studies we reviewed to assess for this protocol did not have a prospective arm that began preoperatively, instead many were observational and initiated the DVS after surgery.

The incidence, which we found (9%) is lower than the mean incidence of DVT found in various other studies (>15%) (5,23). This may be because all our patients were on standard prophylaxis with heparin or Lovenox from postoperative day one and underwent physical therapy.

None of our patients developed symptomatic lower extremity DVT and this may be due to early detection and treatment of subclinical DVT by DVS. None of the patients who had a positive DVS screen developed pulmonary embolism. This again could be attributed to timely detection and treatment of subclinical DVT. Interestingly the only patient who developed pulmonary embolism had a negative DVS for DVT even though his admission was associated with prolong postoperative stay for persistent hypoxia, which eventually revealed bilateral subsegmental PE on CT angiogram of chest on POD 9. This patient had no other known risk factors for VTE and had an uneventful intra-operative course with no blood transfusion requirement. Only 3 out of 124 patients (2.4%) developed bleeding requiring reoperation which is consistent with the bleeding incidence post cardiac surgery in the existing literature, which supports the safety of DVT prophylaxis with heparin or Lovenox in these patients.

Most of the DVTs were found in the first 5 days after surgery as one might expect more as a consequence of this being the most thrombogenic period. We found a similar preponderance of DVT in the ipsilateral site in off pump CABG patients as were seen in other cardiac surgery patients (8).

The higher incidence of DVT in patients who received blood transfusion (54.5%) in the perioperative period was found to be statistically significant p value (<0.05) in our study. This is consistent with prior studies in patients undergoing cardiac surgeries (6,24). Blood transfusion has been established as an important risk factor for the development of DVT due to its pro-inflammatory effect as well as those patients who had bleeding disorders prompting the decision to withhold DVT prophylaxis despite mechanical (non-pharmacological) thromboprophylaxis (25).

An article based on a national database found the incidence of DVT to be 2% in cardiac surgery patients, but this was a retrospective study, and it did not distinguish symptomatic from asymptomatic DVT (3).

The paucity of on pump CABG patients precluded a suitable comparison between the two with respect to DVT. There was no statistically significant difference in the DVT rates between valvular and CABG surgeries.

With general surgery patients, risk factors associated with significantly increased risk of postoperative DVT were operative time more than 4 hours (odds ratio, 2.72), need for blood transfusion (odds ratio, 2.3), and postoperative cardiac arrest. Previously published studies on this topic have similarly identified these factors to be associated with postoperative DVT. Our study re-enforces the importance of identifying patients who are at high risk of developing DVT post cardiac surgery and ensuring adequate DVT prophylaxis for such patients.

We did find higher DVT rates among active smokers and patients with reintubation, but this did not achieve statistical significance. Larger sample size studies are needed to confirm this.

## Limitations

Our cohort of patients was small, and the recruitment process was by consecutive enrollment without randomization. Our center’s minimally invasive approach to isolated valves limited our sample population from undergoing subgroup analysis. Patients unable to return for 30-day follow-up scan were excluded. We did not assess the dose response effect of blood transfusion that might have helped us to establish blood transfusion as a cause of DVT.

## CONCLUSION

Despite optimal thromboprophylaxis and physical therapy, 9% of cardiac surgery patients may develop asymptomatic DVT. Despite limited sample size, we found the DVT group included a higher percentage of patients who smoked, required blood transfusion, and experienced prolonged intubation or reintubation.

## Disclosure

JTM discloses a financial relationship with Medtronic, Inc. Other authors do not have any disclosures.

## Data Availability

All data produced in the present work are contained in the manuscript.

## Acknowledgments

The authorship is based on first-last-author-emphasis. We thank **Ty** and **Oksana Popova**, and other Vascular Technologists in the Vascular Department of Radiology for assistance in receiving and performing study venous duplex studies. We thank the Department of Research at Staten Island University Hospital, Northwell Health (Staten Island, NY) for allowing support and regulatory guidance: **Maria Scibilia RN** and **Meagan Sills MPH**. In addition, we thank the volunteers who assisted with data collection **(Christopher Aseervatham, Carman Chan, Yakov Groysman).** Special thanks to **Diane Johnston** for assisting with patient registration and coding for study scans.

